# Higher Population Coverage with Typhoid Conjugate Vaccine is Needed to Induce Herd Protection: Evidence from a Cluster-Randomized Trial in Urban Bangladesh

**DOI:** 10.64898/2026.06.08.26355125

**Authors:** Faisal Ahmmed, Farhana Khanam, Md Taufiqul Islam, Se Eun Park, Beatrice Ongadi, Justin Im, Yiyuan Zhang, Ashraful Islam Khan, Asma Binte Aziz, Afroza Akter, Benjamin Ngugi, Cecilia Kathure Mbae, Dilruba Nasrin, Edlawit Mesfin Getach, Fahima Chowdhury, Joo-Young Esther Lee, Kassa Haile, Kelvin Kering, Moses Mwangi, Martin Bundi, Meseret Gebre Behute, Md Golam Firoj, Suman Kanungo, Suneth Agampodi, Sadia Isfat Ara Rahman, Xinxue Liu, Andrew J Pollard, K Zaman, Samuel Kariuki, Firdausi Qadri, John D Clemens

**Affiliations:** International Centre for Diarrhoeal Disease Research, Bangladesh (icddr,b), Dhaka, Bangladesh; International Vaccine Institute, Seoul, Republic of Korea; Yonsei University Graduate School of Public Health, Seoul, Republic of Korea; Kenya Medical Research Institute (KEMRI), Nairobi, Kenya; Research Investment for Global Health Technology (RIGHT) Foundation, Seoul, Republic of Korea; Oxford Vaccine Group, Department of Paediatrics, University of Oxford, and the NIHR Oxford Biomedical Research Centre, Oxford, UK; Department of Medicine, Center for Vaccine Development and Global Health, University of Maryland School of Medicine, Baltimore, MD, US; Armauer Hansen Research Institute, Addis Ababa, Ethiopia; ICMR - National Institute of Cholera and Enteric Diseases, Kolkata, West Bengal, India; UCLA Fielding School of Public Health, Los Angeles, CA 90095-1772, USA

## Abstract

**Introduction:** A cluster randomized trial (CRT) in Bangladesh found that Vi-tetanus toxoid (Vi-TT) vaccine conferred 85% protection to vaccinees at 18 months of follow-up; however, it failed to confer significant herd protection to non-vaccinees.

**Methods:** In the CRT, children aged 9 months to <16 years from 150 clusters received one dose of either Vi-TT or Japanese encephalitis vaccine. To evaluate whether herd protection was evident, we analyzed two years of follow-up data by quartiles of clusters with ascending levels of Vi-TT coverage.

**Results:** The average vaccine coverage across all clusters was 69% among targeted children and 20% in the entire population. Vi-TT provided 83% (95%CI: 74%,89%) total protection among vaccinees, 12% (95%CI: -17%,34%) indirect protection among non-vaccinees, and 53% (95%CI: 40%,63%) overall protection among entire population, irrespective of vaccine coverage. Significant herd protection (47%; 95%CI: 3%,71%) was observed only in the highest quartile of vaccine coverage in the entire population (range 21.5%-26%). No herd protection was evident among targeted children (<16 years), including the highest coverage quartile (72.1%-78.9%), where protection was - 8% (95%CI: -108%,44%).

**Conclusion:** Vaccine herd protection, essential for typhoid conjugate vaccines to achieve high overall protection, was evident only in clusters with higher, albeit still modest, coverage in the entire population. Since even high coverage among children <16 years did not generate significant herd protection, higher vaccine coverage across all age groups, including vaccination of adults, may be necessary for TCVs to confer the combined direct and herd protection required for effective typhoid control.

**Summary box:** *What is already known on this topic?:* Vi-tetanus toxoid (Vi-TT) vaccine provides high-level protection against typhoid fever in vaccinated children. However, overall analysis of a cluster-randomized trial of Vi-TT given to children under 16 years of age found no evidence of indirect (herd) protection by Vi-TT to unvaccinated individuals.

*What this study adds:* In further analysis of the cluster randomized trial of Vi-TT, significant indirect protection (47%; 95%CI:3%,71%) was observed, but only in clusters with the highest levels of vaccine coverage of the entire population. In contrast, we found no evidence of vaccine herd protection in clusters with the highest levels of vaccine coverage of children, who were targeted by vaccination.

*How this study might affect research, practice, or policy:*

- Achieving herd protection from typhoid conjugate vaccines (TCVs) may require higher vaccine coverage across the entire population, not only among children. Vaccination strategies that include adults may be necessary to maximize the public health impact of TCVs.
- These findings have important implications for typhoid control policies in endemic settings.

## Introduction

Typhoid fever, caused by *Salmonella enterica* serotype Typhi (*Salmonella* Typhi), remains a pressing public health concern. In 2019, there were an estimated nine million cases of typhoid fever, leading to 110,000 deaths globally, particularly in low- and middle-income countries in Asia and Africa [1]. As *Salmonella* Typhi exclusively infects humans, typhoid elimination may be possible through effective control measures. The World Health Organization (WHO) has recommended the introduction of typhoid conjugate vaccines (TCVs) in national childhood immunization programmes of high typhoid burden countries, accompanied by catch-up vaccination campaigns for children aged 6 months to <16 years [2]. In addition to vaccination, health education, improvements in water quality and better sanitation practices are important components of a comprehensive strategy to eliminate typhoid [3-6].

Clinical trials with a WHO-prequalified Vi-TT (Vi-polysaccharide conjugated to tetanus toxoid) vaccine administered to children aged 9 months to <16 years in Nepal, Malawi, and Bangladesh demonstrated significant protection among vaccinees, ranging from 79% to 85% [7-12]. The cluster-randomized trial (CRT), carried out in an urban setting in Dhaka, Bangladesh, demonstrated 85% total protection (95%CI: 76%,91%) of vaccine recipients at 18 months post-vaccination, but no significant indirect protection (19%; 95%CI: -12%,41%) of unvaccinated individuals. To further investigate the surprising absence of indirect protection by Vi-TT, we earlier investigated whether this absence could have resulted from ingress of typhoid into the clusters from the outside. To evaluate this possibility, we used a “fried-egg” analysis that focused only on the inner core of the populations in the clusters. However, this approach also did not show significant herd effects, with indirect protection estimated at -42% (P = 0.25) among unvaccinated participants in the innermost 25% of the clusters [17].

The failure of Vi-TT to confer significant indirect protection in Bangladesh contrasted with an earlier CRT of Vi-solysaccharide (Vi-PS) in which children and adults aged 2 years and older were vaccinated, and significant indirect protection of 44% (95%CI: 2%,69%) was observed [13, 14]. Yet, similar to the Bangladesh trial of Vi-TT vaccine, no indirect protection (–10%; 95%CI: –116%,44%) was observed when Vi-PS was administered only to 2-16 years children in another CRT in Karachi [15].

The results from Kolkata and Karachi suggest an alternative explanation for low level of indirect protection in the Bangladesh trial: not to vaccinate adults along with children, resulting in levels of V-TT coverage of the population that were insufficient to interrupt transmission of typhoid. The cluster-randomized design of the Bangladesh trial allowed to examine whether the subset of clusters with higher levels of Vi-TT coverage exhibited indirect protection. Herein, we report the results of this reanalysis.

## Methods

### Description of the trial

As described elsewhere, a participant and observer-blind CRT was conducted in wards 2, 3, and 5 of Mirpur, a densely populated urban area of Dhaka, Bangladesh [9]. Prior to vaccination, between 14 February and 25 March 2018, a baseline census was conducted to collect data on participants’ demographics, socioeconomic status, household water, sanitation, hygiene (WASH) conditions and geographic location. The study area, with a baseline population of 205760, was segmented into 150 geographically defined clusters, each comprising approximately 1200 residents. Clusters were randomly assigned to receive either the Vi-TT vaccine or the SA-14-14-2 Japanese encephalitis (JE) vaccine. Randomization was stratified by geographic ward, median distance from the cluster’s center to the adjacent treatment facility, and whether the number of children aged 9 months to <16 years in the cluster was above or below the median. Healthy, non-pregnant individuals 9 months to <16 years who had not received typhoid or JE vaccines in the past three years (although typhoid vaccine had never before been used programmatically or in previous studies in the study area) were offered a single dose of the vaccine assigned to their cluster following acquisition of informed written consent from a parent or guardian. During the baseline round (15 April–15 May 2018), Vi-TT was administered intramuscularly and JE vaccine subcutaneously. Three catch-up vaccination rounds were implemented at roughly six-month intervals: September-December 2018, April-May 2019, and October-November 2019, to include children who became eligible or migrated into the study area after the initial vaccination. Details of the randomization, masking, and vaccine administration process have been published previously [9].

Passive surveillance for typhoid and paratyphoid fever, begin on 26 February 2018 in eight designated treatment facilities, targeting participants with ≥2 days of fever or an axillary temperature ≥38.0°C across the entire population. Following informed written consent, eligible participants were enrolled in passive surveillance with systematic recording of clinical features and collection of 3-5 ml of blood specimens for culture using the BacT/ALERT system and standard microbiological procedures. Treatment was provided by designated study physicians and adjusted as needed based on antibiotic susceptibility test reports [9]. The onset of illness was considered the start date of fever. Febrile visits occurring within 14 days of discharge from the previous febrile visit were considered as a single febrile episode. A typhoid case was defined as a febrile episode with at least one blood culture yielding *Salmonella* Typhi. We further required that a post-discharge household visit verify that the individual whose name had been given in the treatment centre had indeed sought care on the recorded date.

### Statistical analysis

The primary objective was to assess indirect protection conferred by the Vi-TT at varying levels of vaccine coverage of the clusters by comparing the incidence of typhoid fever in unvaccinated individuals in Vi-TT clusters with that among unvaccinated individuals in JE clusters. As secondary objectives, we also measured the two other measures of vaccine herd protection by Vi-TT, total and overall protection, at different levels of coverage. Total protection was defined as the relative reduction in typhoid incidence among Vi-TT recipients compared to JE recipients. Overall protection compared the incidence of typhoid in all participants of Vi-TT clusters versus all participants of JE clusters. The analyses were carried out in a dynamic cohort designed to capture the entire population over the 23-month follow-up period, accounting for the highly mobile nature of the population. This cohort included all persons living in the study clusters at baseline, newborns and in-migrants.

Follow-up began on 30 April 2018 for non-vaccinees (midpoint of the baseline campaign) and on each participant’s vaccination date for vaccinees to assess indirect, total, and overall vaccine protection. For persons entering the population after baseline, the start date was the date of entry into the study clusters. For persons who were initially unvaccinated at either of these two start-dates but later became vaccinated, we included their follow-up using the later start date of vaccination for estimating total vaccine protection. Follow-up was censored by the date of typhoid fever, outmigration from study arm, date of death, or the end of the 23-month surveillance period, whichever occurred first. For analysis of indirect vaccine protection, an additional censoring criterion was the date of vaccination for initially unvaccinated children who later became vaccinated. Counting of typhoid episodes began the day after the start date. Only the first episodes of typhoid were included in the analyses. Sample size justification for the trial is provided in the original report [9].

Vaccine coverage in each cluster was calculated using two strategies: one based on the entire population (“entire population coverage”) and the other on the population 9 months to <16 years of age (“<16-year coverage”); both were assessed from the start date used for analysis of overall vaccine protection, and age denoted age at the start date. To calculate this, we divided the person-time of the vaccinated participants after vaccination by the total person-time of either all residents or residents aged 9 months to <16 years, followed from the start date in the clusters, censoring follow-up as already described. For each type of coverage, we stratified clusters into four quartiles of ascending coverage.

Baseline characteristics of participants were compared between two study arms within the different vaccine coverage quartiles. Descriptive statistics were used to summarize data, with categorical variables presented as frequency and percentages, and continuous variables presented as mean with standard deviation. To assess differences between the two arms, generalized linear mixed-effect models with log link function were used for analysis of binary response variables, and linear mixed-effects models were used for continuous response variables. These models accounted for the design effect of clusters by treating each cluster as a random effect.

Within each coverage quartile, the incidence of typhoid fever in Vi-TT clusters was compared to that in JE clusters to calculate indirect, total, and overall protection. To estimate hazard ratios (HRs) that compared rates of typhoid in the Vi-TT and JE arms, while accounting for the design effect of cluster randomization as well as prespecified baseline covariates, we exponentiated the vaccine arm variable in Cox proportional hazard models in which robust variance was calculated considering each cluster as a group. We conducted subgroup analyses in different age groups (under 16 years and ≥16 years at the start date) using the same Cox proportional hazard models. We applied the same vaccine coverage ranges for quartiles, defined separately entire population coverage and <16-year coverage, for all analyses. Stratifying variables for randomization were forced as independent variables in models. Additional baseline covariates, age at start date, sex, presence of a flush toilet, access to a safe drinking water source, use of treated drinking water, and hand washing habits after defecation were included if they differed between two study arms at P<0.05, a parsimonious strategy to prevent overfitting of the models. Vaccine protection was calculated as (1-HR) × 100%. A two-tailed *P* value of <0.05 was considered statistically significant. All statistical analyses were conducted using R software version 4.2.1 (2022-06-23 ucrt) [16].

## Results

### Assembly of the study population

The baseline population was 205,760 persons, with 114,925 in-migrants, 6,109 births, and 1,829 deaths occurring during the 23 months of follow-up. At the end of the follow-up period, person-time was considered for 163,371 and 163,423 participants for Vi-TT and JE arms, respectively. After excluding 181 ineligible typhoid fever episodes, 655 typhoid fever episodes were included in the analysis (Figure 1).

**Figure 1.**
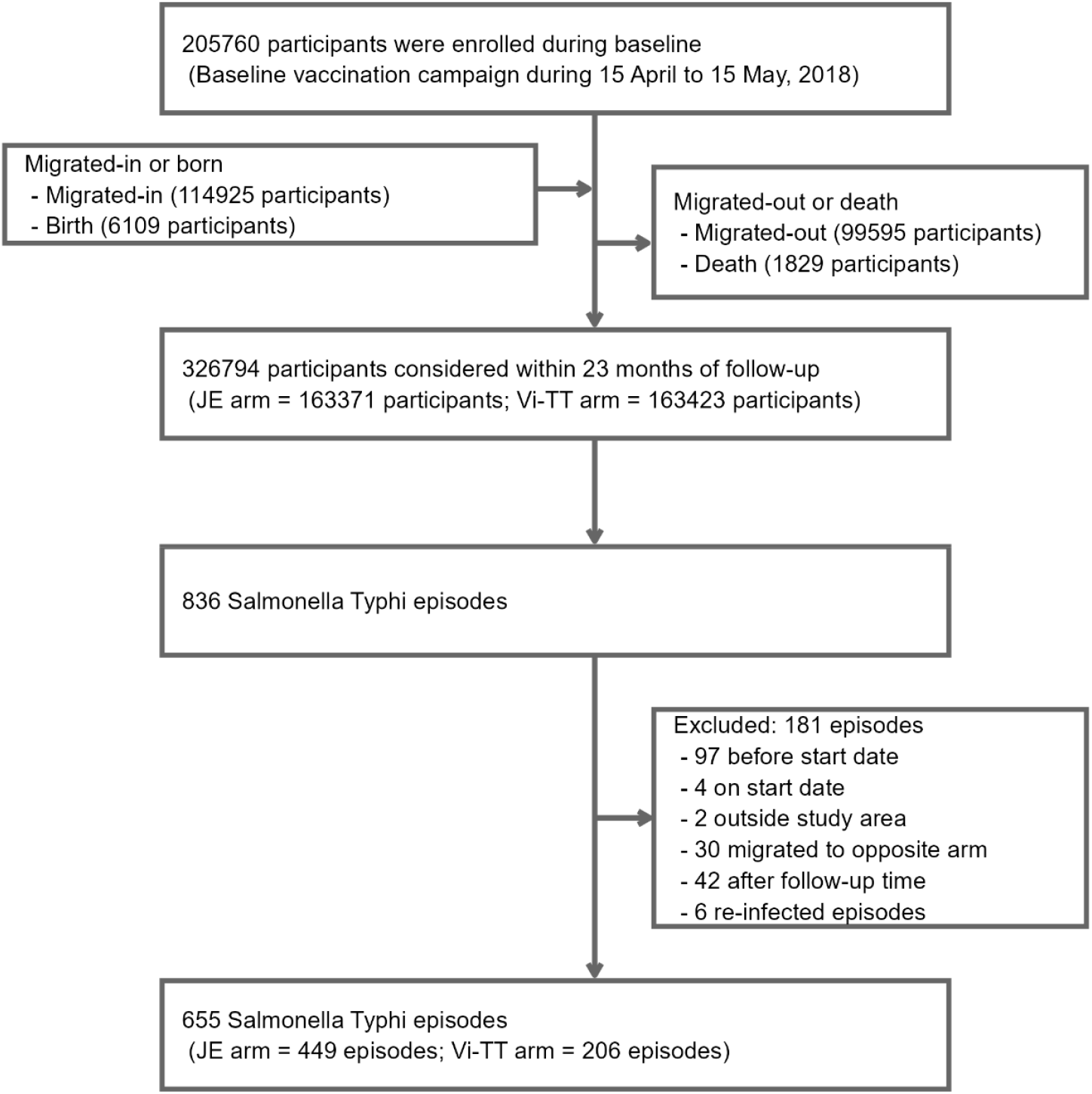
CONSORT for assembly of study participants for analysis.

### Vaccine coverage in the clusters

Figure 2 shows the relationship between the entire population vaccine coverage and <16-year coverage in the clusters of the Vi-TT and JE arms. The average vaccine coverage of all clusters was 69% for <16-year coverage and 20% for the entire population coverage. As expected, there was a linear relationship between the entire population coverage and <16-year coverage. However, there was an appreciable splay from a strict linear relationship, indicating that separate evaluation of Vi-TT protection by the two different types of coverage might be possible.

**Figure 2.**
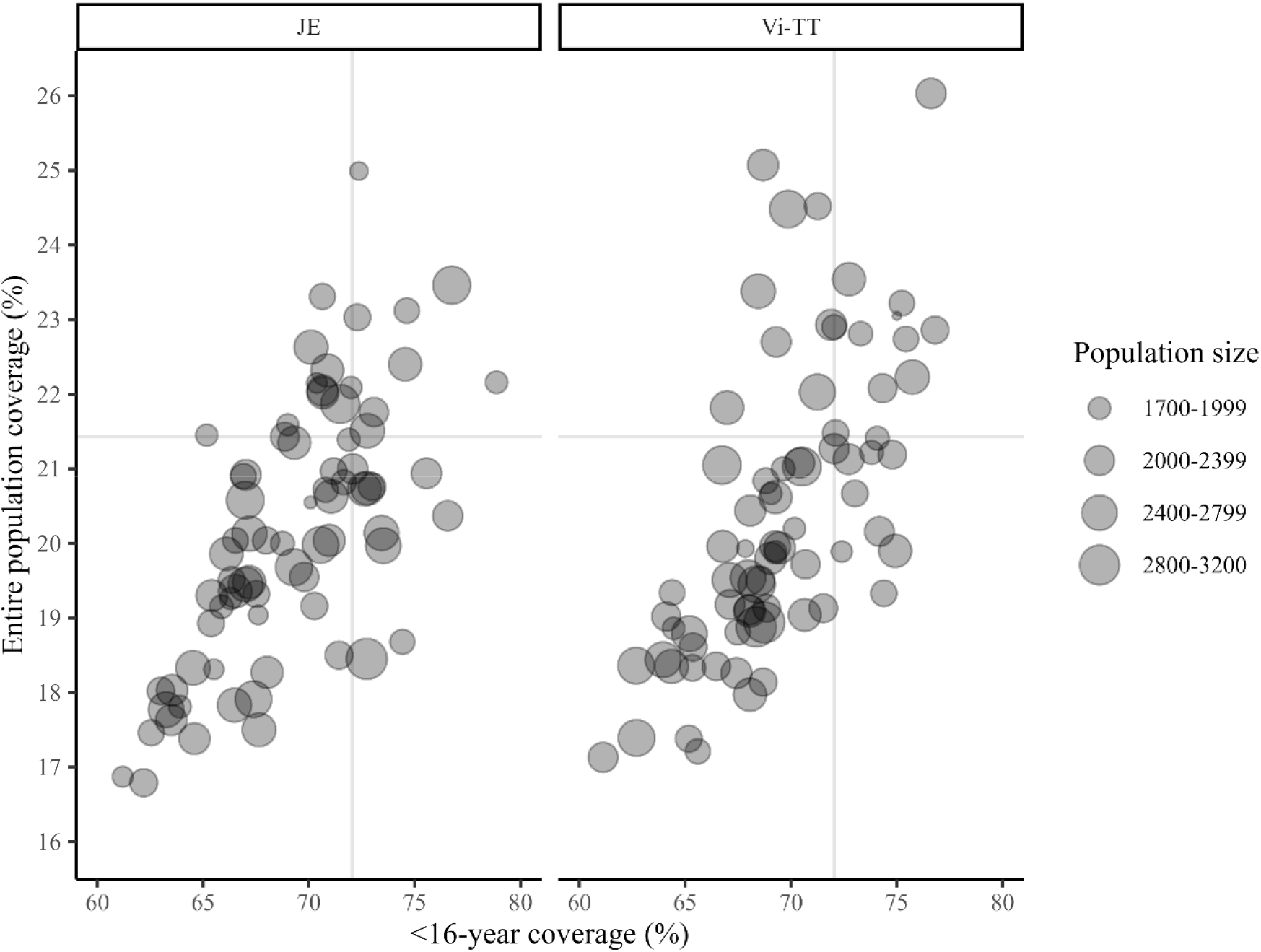
Relationship between entire population vaccine coverage and vaccine coverage of children under 16 years of age. **Legend:** The X-axis represents under 16-year vaccine coverage (ranging from 61.1% to 78.9%), while the Y-axis represents population level vaccine coverage (ranging from 16.8% to 26.0%). Each point indicates the study participants in a cluster, with population size distinguishing four groups: 1700-1999, 2000-2399, 2400-2799, and 2800-3200 participants. Vertical and horizontal lines represent the third quartile of vaccine coverage.

### Baseline comparability by type of coverage

The ranges of entire population coverage for the quartiles were 16.8% to 18.9%; 19.0% to 20.0%; 20.1% to 21.4% and 21.5% to 26.0%. Quartiles for <16-year vaccine coverage were 61.1% to 66.9%; 67.0% to 69.3%; 69.4% to 72.0%; and 72.1% to 78.9%. Supplementary Tables 1-6 depict JE and Vi-TT arms for several baseline variables, by level of vaccine coverage, by type of vaccine protection evaluated, and by age group evaluated for vaccine protection. The trial arms were generally similar, though some significant differences were noted for age, sex, presence of flush toilet in the house, use of treated drinking water, distance of midpoint of cluster to the nearest study clinic, and study ward of residence, depending on the analysis.

### Indirect protection by vaccination with Vi-TT vaccine

Table 1 shows indirect protection of non-vaccinees by Vi-TT for each quartile of coverage of the entire population. In the highest coverage quartile (21.5% to 26.0%), indirect protection was 47% (95% CI: 3%,71%; *P*=0.041). In other coverage quartiles, there was no evidence of indirect protection. When evaluated by <16-year coverage there was no evidence of indirect protection, even in the highest coverage quartile (72.1% to 78.9%).

**Table 1.**
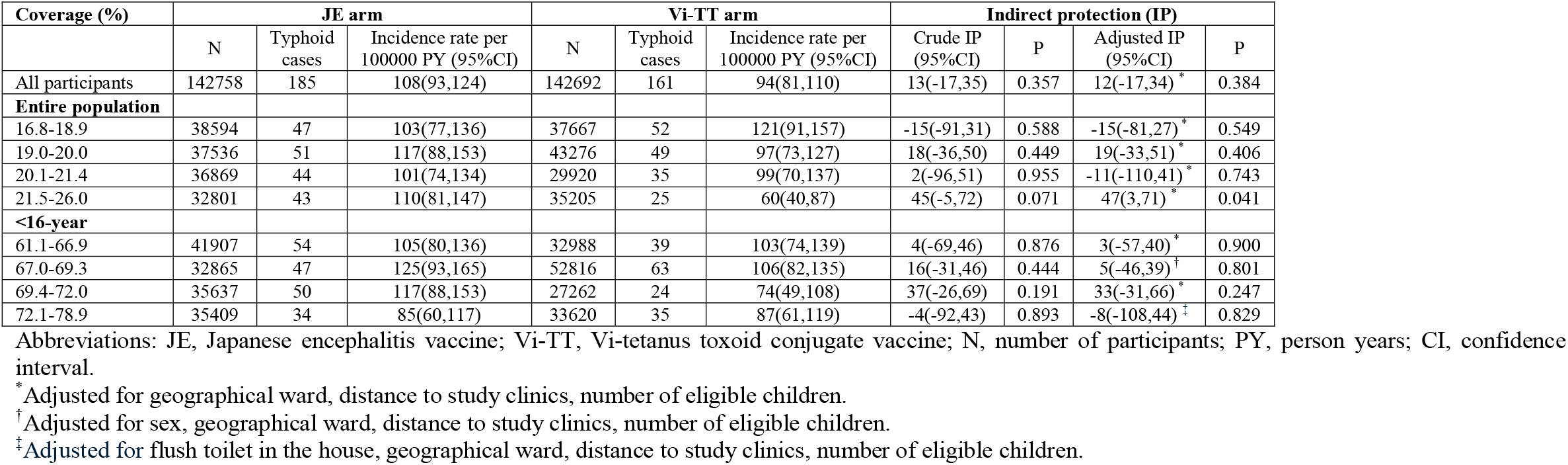
Indirect protection by Vi-TT vaccine.

Among children under 16 years at the start date, there was no evidence of indirect protection in the different quartiles for coverage of either the entire population or < 16-year-olds (Table 2). In contrast, indirect protection increased as vaccine coverage of the entire population increased in ≥ 16-year-olds, reaching 58% (95%CI: 18%,78%; *P*=0.011) in the highest quartile of coverage. Indirect protection increased somewhat with increasing coverage of < 16-year-olds, though it did not reach statistical significance (Table 3).

**Table 2.** Indirect protection by Vi-TT vaccine among non-vaccinees under 16 years of age.

| Coverage (%) | JE arm |  |  | Vi-TT arm |  |  | Indirect protection (IP) |  |  |  |
| --- | --- | --- | --- | --- | --- | --- | --- | --- | --- | --- |
|  | N | Typhoid cases | Incidence rate per 100000 PY (95%CI) | N | Typhoid cases | Incidence rate per 100000 PY (95%CI) | Crude IP (95%CI) | P | Adjusted IP* (95%CI) | P |
| All participants | 31272 | 92 | 363(294,443) | 31606 | 86 | 340(274,418) | 7(-36,36) | 0.705 | 4(-38,33) | 0.831 |
| Entire population |  |  |  |  |  |  |  |  |  |  |
| 16.8-18.9 | 8156 | 27 | 405(273,581) | 8127 | 23 | 351(228,517) | 14(-52,51) | 0.604 | 15(-44,50) | 0.553 |
| 19.0-20.0 | 8194 | 28 | 432(293,615) | 9217 | 23 | 319(208,470) | 28(-37,62) | 0.320 | 28(-29,60) | 0.263 |
| 20.1-21.4 | 7986 | 20 | 314(198,476) | 6547 | 27 | 526(354,754) | -65(-279,28) | 0.236 | -95(-326,10) | 0.092 |
| 21.5-26.0 | 7351 | 17 | 291(176,456) | 8185 | 13 | 204(114,339) | 30(-86,73) | 0.477 | 30(-68,71) | 0.422 |
| <16-year |  |  |  |  |  |  |  |  |  |  |
| 61.1-66.9 | 9236 | 31 | 382(265,535) | 7418 | 20 | 331(209,501) | 16(-59,55) | 0.598 | 12(-56,50) | 0.663 |
| 67.0-69.3 | 7398 | 21 | 367(234,551) | 11822 | 29 | 326(223,462) | 12(-70,55) | 0.701 | -13(-101,37) | 0.679 |
| 69.4-72.0 | 7690 | 29 | 462(316,655) | 6104 | 14 | 285(163,465) | 38(-25,69) | 0.182 | 37(-16,66) | 0.138 |
| 72.1-78.9 | 7340 | 11 | 210(111,363) | 6845 | 23 | 424(276,626) | -109(-385,10) | 0.088 | -89(-368,24) | 0.171 |

Abbreviations: JE, Japanese encephalitis vaccine; Vi-TT, Vi-tetanus toxoid conjugate vaccine; N, number of participants; PY, person years; CI, confidence interval.
\*Adjusted for geographical ward, distance to study clinics, number of eligible children.

**Table 3.** Indirect protection by Vi-TT vaccine among non-vaccinees ≥16 years of age.

| Coverage (%) | JE arm |  |  | Vi-TT arm |  |  | Indirect protection (IP) |  |  |  |
| --- | --- | --- | --- | --- | --- | --- | --- | --- | --- | --- |
|  | N | Typhoid cases | Incidence rate per 100000 PY (95%CI) | N | Typhoid cases | Incidence rate per 100000 PY (95%CI) | Crude IP (95%CI) | P | Adjusted IP* (95%CI) | P |
| All participants | 111601 | 93 | 64(52,77) | 111207 | 75 | 52(41,64) | 19(-14,43) | 0.222 | 19(-13,42) | 0.211 |
| **Entire population** |  |  |  |  |  |  |  |  |  |  |
| 16.8-18.9 | 30456 | 20 | 52(32,78) | 29556 | 29 | 79(54,112) | -52(-176,16) | 0.171 | -50(-149,10) | 0.117 |
| 19.0-20.0 | 29365 | 23 | 62(41,92) | 34073 | 26 | 60(40,87) | 4(-83,49) | 0.908 | 5(-80,50) | 0.872 |
| 20.1-21.4 | 28907 | 24 | 64(42,94) | 23387 | 8 | 27(13,50) | 59(5,82) | 0.037 | 53(-3,79) | 0.059 |
| 21.5-26.0 | 25477 | 26 | 78(52,113) | 27065 | 12 | 34(19,58) | 56(17,77) | 0.012 | 58(18,78) | 0.011 |
| **<16-year** |  |  |  |  |  |  |  |  |  |  |
| 61.1-66.9 | 32697 | 23 | 53(35,78) | 25583 | 19 | 59(37,91) | -10(-116,44) | 0.777 | -8(-91,40) | 0.805 |
| 67.0-69.3 | 25484 | 26 | 82(55,118) | 41020 | 34 | 67(47,93) | 18(-44,53) | 0.484 | 15(-56,53) | 0.606 |
| 69.4-72.0 | 27974 | 21 | 58(37,86) | 21177 | 10 | 36(19,64) | 37(-60,75) | 0.329 | 28(-87,72) | 0.498 |
| 72.1-78.9 | 28092 | 23 | 66(43,98) | 26801 | 12 | 34(19,58) | 47(-5,73) | 0.067 | 35(-38,69) | 0.266 |

Abbreviations: JE, Japanese encephalitis vaccine; Vi-TT, Vi-tetanus toxoid conjugate vaccine; N, number of participants; PY, person years; CI, confidence interval.
\*Adjusted for geographical ward, distance to study clinics, number of eligible children

### Overall and Total Protection by vaccination with Vi-TT vaccine

Overall protection of the entire population provided by the Vi-TT vaccine increased as vaccine coverage of the entire population increased, rising to 66% (95%CI: 45%, 78%; *P*<0.001) in the highest quartile of coverage, but there was no consistent trend when assessed by coverage of < 16-year-olds (Supplementary Table 7). There was no increase in overall protection by quartile of either < 16-year-olds or the entire population among < 16-year-olds (Supplementary Table 9). Analyses of overall protection of ≥ 16-year-olds were identical to analyses of indirect vaccine protection in this age group, since this age group was not vaccinated. Total protection remained consistent across all quartiles when stratified by quartile of coverage of either the entire population or < 16-year-olds (Supplementary Table 8). Age-stratification for assessment of total protection was not meaningful, as only children <16 years of age were vaccinated.

## Discussion

Our earlier published analysis of the Vi-TT CRT in Bangladesh did not reveal significant herd protection overall, nor in a ‘fried-egg’ analysis, designed to minimize the attenuation of measured indirect protection owing to transmission of typhoid into the clusters from the outside [9, 17]. This was surprising, as an earlier generation, less protective typhoid vaccine, Vi-PS, given to all persons aged 2 years and above, had shown clear evidence of vaccine herd protection when tested in a CRT in Kolkata. Among several possible explanations, this paper evaluated whether restricting Vi-TT vaccination to children resulted in insufficient of vaccine coverage in the entire population to substantially interrupt typhoid transmission.

To evaluate this possibility, we took advantage of the inter-cluster variability of vaccine coverage in the Bangladesh trial and evaluated measures of Vi-TT herd protection by the level of vaccine coverage. Our findings show that clusters with the highest levels of vaccine coverage of the entire population, in the range of 21.5% to 26.0%, exhibited evidence of both augmented indirect and overall vaccine protection in comparison with clusters with lower coverage. However, when evaluated not by vaccine coverage of the entire population but by coverage of < 16-year-olds, the age group targeted for Vi-TT, vaccine herd protection was not evident even in the highest quartile. Interestingly, the vaccine herd protection that became evident as vaccine coverage of the entire population increased was observed in persons 16 years and older, but not in younger persons.

Separate examination of relationships for Vi-TT coverage of the entire population and coverage of < 16-year-olds was possible because of the divergence between these two types of coverage in many clusters (Figure 2). A major finding was that the level of vaccine coverage of the entire population, but not of children <16 years of age, was the key determinant of Vi-TT herd protection, suggesting that children may not be the main source of transmission of typhoid in the population under study. This may seem counterintuitive, since rates of typhoid disease seen in treatment facilities were considerably higher in children than in adults in our study population. However, a previous disease burden study conducted in the same population reported a higher sero-incidence of typhoid infection in adults than children, indicating that adults could be a major source of infection [18]. Moreover, the explanation may lie in the fact that food, a major vehicle of typhoid transmission, is more commonly prepared by adults than by children in this setting [19, 20]. As well, chronic fecal excretion of typhoid is seen primarily in adults, and although vaccination of chronic excretors does not halt excretion, it does prevent the initial infections that lead to chronic excretion [21].

Vaccine herd effects were seen only in persons ≥16 years. Although the mechanism for this observation remains to be elucidated, it could have resulted from ingestion of a higher inoculum of typhoid organisms in children than adults or from the higher level of natural immunity to typhoid in adults than in children. The modest reduction in typhoid transmission from low levels of vaccine coverage of the entire population, combined with other factors, likely explains the age-dependent herd effects observed in our analyses. If true, a greater reduction of transmission achieved by higher levels of vaccine coverage might be expected to result in herd benefits for both children and adults. Whatever the explanation, the data indicate that substantial increases in the levels of vaccine coverage of the entire population may be needed to achieve major levels of typhoid control when TCVs are implemented in practice. Indeed, the overall reduction of typhoid conferred by Vi-TT in the current analysis of the Bangladesh trial was only 53% despite an intensive program of childhood vaccination with multiple catch-up rounds. Conversely, the potential of a moderately effective enteric vaccine to yield substantial herd and overall protection is illustrated by killed oral cholera vaccines, which, despite conferring approximately 65% direct protection against cholera, were found in a modeling study to reduce the risk of cholera in unvaccinated persons by 89% despite a vaccine coverage of only 50% [22]. These findings suggest that future modeling work could help estimate the level of TCV coverage needed to achieve herd protection against typhoid. Such models could also clarify whether vaccinating adults is important for reducing transmission.

Our study has limitations. Firstly, this was a post hoc analysis. Secondly, our study was conducted in a densely populated urban slum in Dhaka, where the force of infection of typhoid is higher than in many populations with endemic typhoid. Whether our findings would be replicated in lower incidence settings is not certain. Thirdly, our conclusions are based on a program of vaccination of only children. The effects of higher vaccine coverage attained by vaccination of both children and adults remain to be demonstrated.

In conclusion, the main published results of the trial failed to demonstrate a significant level of herd protection by vaccination of children with Vi-TT, with the result that vaccination had only a modest impact in reducing the total burden of typhoid in the population [9]. However, these overall results concealed the fact that herd protection was evident in clusters with higher vaccine coverage of the entire population. The only feasible way of achieving high levels of vaccine coverage of the entire population in practice will be to vaccinate both children and adults. This broader age-targeting strategy, however, will require a major revision of the current WHO recommendation to vaccinate only children with TCVs. Before making such a policy revision, it will be essential to evaluate in future studies the hypothesis that combined vaccination of children and adults will result in substantial vaccine herd protection and a major reduction of a population’s total typhoid burden.

## Data Availability

A de-identified analytical data set will be made available upon requests directed to the Institutional Review Board (IRB) coordinator of the icddr,b at. Only after approval of a proposal data can be shared through a secure online platform. Approval of the proposal will be subject to scientific review by the IRB at icddr,b. Sharing of data will also be subject to the published data access rules of the icddr,b. The requestor will need to sign a standard data access agreement required by the icddr,b.

## Author Contribution

JDC, FQ, and SK conceptualized the study protocol and acquired funding. Project administration and supervision was undertaken by JDC, FQ, SK, and FK. FA, JDC, and FQ developed the methodology. FA and FK curated the data, with FA responsible for developing the code of the statistical software. Formal analysis and visualization were conducted by FA, FK, MTI, SEP, BO, JI, and YZ and drafted the original manuscript. Result validation was performed by JDC, FQ, SK, and KZ. All authors reviewed the manuscript, and FA, FK, MTI, SEP, BO, JI, and YZ revised it further under the scientific guidance of JDC, FQ, SK, and KZ. All authors approved the final version of the manuscript and agree to be accountable for all aspects of the work.

## Ethical approval

The study protocol was approved by the institutional review boards of the icddr,b, University of Oxford and University of Maryland. The trial is registered under ISRCTN11643110.

## Acknowledgements

The Extended Analysis of Archival Datasets Project team is grateful to the Bill and Melinda Gates Foundation for their support to this project. The icddr,b, IVI, KEMRI and AHRI are grateful to their core donors (icddr,b: Governments of Bangladesh and Canada; IVI: Governments of Korea, Sweden, India, Finland, Denmark, the Philippines and Thailand; KEMRI: Ministry of Health; AHRI: Federal Ministry of Health of Ethiopia and the AHRI core fund from SIDA and NORAD;) for providing unrestricted support. We sincerely thank the large number of research staff who contributed to the original trial.

