## Supplementary tables 1-9 for "Higher Population Coverage with Typhoid Conjugate Vaccine is Needed to Induce Herd Protection: Evidence from a Cluster-Randomized Trial in Urban Bangladesh"

**Supplementary table 1. Baseline characteristics of non-vaccinees in the two study arms, stratified by vaccine coverage of the entire population**

| **Characteristic** | **Coverage: 16.8% to 18.9%** | | | **Coverage: 19.0% to 20.0%** | | | **Coverage: 20.1% to 21.4%** | | | **Coverage: 21.5% to 26.0%** | | |
| --- | --- | --- | --- | --- | --- | --- | --- | --- | --- | --- | --- | --- |
|  | JE  (n=37959) | Vi-TT (n=36829) | P^*^ | JE  (n=31187) | Vi-TT  (n=42351) | P^*^ | JE  (n=41341) | Vi-TT  (n=29026) | P^*^ | JE  (n=32271) | Vi-TT  (n=34486) | P^*^ |
| Mean age, year at start date | 28.2 ± 16.8 | 28 ± 16.8 | 0.599 | 28.0 ± 16.8 | 28.2 ± 16.9 | 0.442 | 28.1 ± 17.1 | 27.8 ± 16.9 | 0.367 | 27.6 ± 16.9 | 27.4 ± 16.9 | 0.571 |
| Number of male participants | 18838(49.6) | 18118(49.2) | 0.400 | 18104(49.6) | 21023(49.6) | 0.864 | 17764(49.4) | 14559(50.2) | 0.147 | 16147(50.0) | 17230(50.0) | 0.893 |
| Flush toilet facility in the house | 3090(8.1) | 2988(8.1) | 0.771 | 2396(6.6) | 3725(8.8) | 0.954 | 1193(3.3) | 1109(3.8) | 0.554 | 569(1.8) | 322(0.9) | 0.285 |
| Own source of drinking water in the house | 16900(44.5) | 16911(45.9) | 0.938 | 15204(41.6) | 18138(42.8) | 0.767 | 12838(35.7) | 10136(34.9) | 0.760 | 11098(34.4) | 9956(28.9) | 0.209 |
| Treated drinking water | 32899(86.7) | 31940(86.7) | 0.575 | 31101(85.1) | 36607(86.4) | 0.878 | 25321(70.3) | 21062(72.6) | 0.391 | 18292(56.7) | 15681(45.5) | 0.383 |
| Handwashing after defecation | 37153(97.9) | 36269(98.5) | 0.530 | 35818(98.0) | 41672(98.4) | 0.790 |  | 28610(98.6) | 0.499 | 30722(95.2) | 32706(94.8) | 0.972 |
| Longer than median distance to the midpoint of nearest treatment facility | 16551(43.6) | 16379(44.5) | 0.964 | 11758(32.2) | 20085(47.4) | 0.701 | 19108(53.1) | 15358(52.9) | 0.950 | 26233(81.3) | 21137(61.3) | 0.171 |
| Number of eligible children at baseline in cluster (above median) | 11374(30.0) | 15586(42.3) | 0.772 | 14990(41.0) | 14210(33.6) | 0.884 | 22108(61.4) | 15235(52.5) | 0.576 | 25179(78.0) | 29817(86.5) | 0.601 |
| Ward of residence |  |  |  |  |  |  |  |  |  |  |  |  |
| 2 | 13031(34.3) | 17375(47.2) | Ref. | 18746(51.3) | 14311(33.8) | Ref. | 17021(47.3) | 10783(37.1) | Ref. | 6280(19.5) | 15213(44.1) | Ref. |
| 3 | 16600(43.7) | 13556(36.8) | 0.234 | 5639(15.4) | 9611(22.7) | 0.667 | 5903(16.4) | 10208(35.2) | 0.661 | 5825(18.1) | 5600(16.2) | 0.634 |
| 5 | 8328(21.9) | 5898(16.0) | 0.731 | 12149(33.3) | 18429(43.5) | 0.077 | 13070(36.3) | 8035(27.7) | 0.951 | 20166(62.5) | 13673(39.6) | 0.097 |

Data are n (%), mean ± standard deviation.

Abbreviations: JE, Japanese encephalitis vaccine; Vi-TT, Vi-tetanus toxoid conjugate vaccine.

^*^P values were generated using generalized linear mixed-effect or linear mixed-effect models, with each cluster considered as a group.

**Supplementary table 2. Baseline characteristics of non-vaccinees in the two study arms, stratified by vaccine coverage of under 16-year olds**

| **Characteristics** | **Coverage: 61.1% to 66.9%** | | | **Coverage: 67.0% to 69.3%** | | | **Coverage: 69.4% to 72.0%** | | | **Coverage: 72.1% to 78.9%** | | |
| --- | --- | --- | --- | --- | --- | --- | --- | --- | --- | --- | --- | --- |
|  | JE  (n=41237) | Vi-TT  (n=32090) | P^*^ | JE^*^  (n=27705) | Vi-TT^†^  (n=45980) | P^*^ | JE^*^  (n=39122) | Vi-TT^†^  (n=31907) | P^*^ | JE^*^  (n=34694) | Vi-TT^†^  (n=32715) | P^*^ |
| Mean age, year at start date | 28.3 ± 17.2 | 28.1 ± 17.1 | 0.594 | 27.6 ± 16.8 | 27.7 ± 16.8 | 0.653 | 28.0 ± 16.9 | 27.6 ± 16.9 | 0.201 | 27.9 ± 16.6 | 28.1 ± 16.8 | 0.587 |
| Number of male participants | 20571(49.9) | 15848(49.4) | 0.342 | 15746(49.0) | 25622(49.7) | 0.132 | 17319(50.0) | 13173(50.0) | 0.948 | 17217(49.6) | 16287(49.8) | 0.769 |
| Flush toilet facility in the house | 3097(7.5) | 2613(8.1) | 0.345 | 1461(4.5) | 3480(6.8) | 0.667 | 996(2.9) | 1342(5.1) | 0.912 | 1694(4.9) | 709(2.2) | 0.016 |
| Own source of drinking water in the house | 20311(49.3) | 14444(45.0) | 0.375 | 11263(35.0) | 19585(38.0) | 0.930 | 11492(33.1) | 9163(34.8) | 0.473 | 12974(37.4) | 11949(36.5) | 0.748 |
| Treated drinking water | 33376(80.9) | 27025(84.2) | 0.341 | 23030(71.6) | 41708(80.9) | 0.328 | 23773(68.6) | 16125(61.2) | 0.121 | 27434(79.1) | 20432(62.5) | 0.235 |
| Handwashing after defecation | 40165(97.4) | 31647(98.6) | 0.492 | 31160(96.9) | 50853(98.7) | 0.083 | 34079(98.3) | 25461(96.6) | 0.154 | 33958(97.9) | 31296(95.7) | 0.290 |
| Longer than median distance to the midpoint of nearest treatment facility | 19353(46.9) | 14014(43.7) | 0.928 | 14674(45.6) | 32634(63.3) | 0.263 | 23025(66.4) | 13352(50.7) | 0.389 | 16598(47.8) | 12959(39.6) | 0.770 |
| Number of eligible children at baseline in cluster (above median) | 16043(38.9) | 14652(45.7) | 0.872 | 14698(45.7) | 27144(52.7) | 0.829 | 21178(61.1) | 18822(71.4) | 0.402 | 21732(62.6) | 14230(43.5) | 0.065 |
| Ward of residence |  |  |  |  |  |  |  |  |  |  |  |  |
| 2 | 9340(22.6) | 9907(30.9) | Ref. | 13058(40.6) | 17686(34.3) | Ref. | 11007(31.7) | 14929(56.7) | Ref. | 21673(62.5) | 15160(46.3) | Ref. |
| 3 | 20530(49.8) | 18085(56.4) | 0.779 | 5718(17.8) | 15483(30.0) | 0.655 | 4203(12.1) | 176(0.7) | 0.647 | 3516(10.1) | 5231(16.0) | 0.823 |
| 5 | 11367(27.6) | 4098(12.8) | 0.104 | 13383(41.6) | 18369(35.6) | 0.790 | 19458(56.1) | 11244(42.7) | 0.227 | 9505(27.4) | 12324(37.7) | 0.725 |

Data are n (%), mean ± standard deviation.

Abbreviations: JE, Japanese encephalitis vaccine; Vi-TT, Vi-tetanus toxoid conjugate vaccine.

^*^P values were generated using generalized linear mixed-effect or linear mixed-effect models, with each cluster considered as a group.

**Supplementary table 3. Baseline characteristics of overall population in the two study arms stratified by vaccine coverage of the entire population**

| **Characteristics** | **Coverage: 16.8% to 18.9%** | | | **Coverage: 19.0% to 20.0%** | | | **Coverage: 20.1% to 21.4%** | | | **Coverage: 21.5% to 26.0%** | | |
| --- | --- | --- | --- | --- | --- | --- | --- | --- | --- | --- | --- | --- |
|  | JE  (n=42655) | Vi-TT  (n=41366) | P^*^ | JE  (n=35397) | Vi-TT  (n=4856) | P^*^ | JE  (n=47591) | Vi-TT  (n=33391) | P^*^ | JE  (n=37728) | Vi-TT  (n=40610) | P^*^ |
| Mean age, year at start date | 25.9 ± 17.2 | 25.7 ± 17.2 | 0.557 | 25.5 ± 17.2 | 25.7 ± 17.2 | 0.289 | 25.3 ± 17.4 | 25.1 ± 17.2 | 0.360 | 24.7 ± 17.2 | 24.4 ± 17.2 | 0.228 |
| Number of male participants | 21167(49.6) | 20429(49.4) | 0.624 | 20654(49.8) | 23800(49.5) | 0.603 | 20469(49.3) | 16659(49.9) | 0.283 | 18843(49.9) | 20210(49.8) | 0.724 |
| Flush toilet facility in the house | 3343(7.8) | 3205(7.7) | 0.744 | 2568(6.2) | 4022(8.4) | 0.976 | 1283(3.1) | 1234(3.7) | 0.575 | 610(1.6) | 346(0.9) | 0.283 |
| Own source of drinking water in the house | 18581(43.6) | 18809(45.5) | 0.871 | 17086(41.2) | 20351(42.3) | 0.755 | 14573(35.1) | 11549(34.6) | 0.769 | 13021(34.5) | 11821(29.1) | 0.220 |
| Treated drinking water | 36799(86.3) | 35739(86.4) | 0.595 | 35226(84.9) | 41354(86.1) | 0.853 | 29080(70.1) | 24115(72.2) | 0.397 | 21151(56.1) | 18321(45.1) | 0.392 |
| Handwashing after defecation | 41724(97.8) | 40707(98.4) | 0.583 | 40637(97.9) | 47308(98.4) | 0.702 | 41128(99.1) | 32917(98.6) | 0.454 | 35899(95.2) | 38482(94.8) | 0.987 |
| Longer than median distance to the midpoint of nearest treatment facility | 18553(43.5) | 18403(44.5) | 0.964 | 13436(32.4) | 22746(47.3) | 0.704 | 22016(53.1) | 17511(52.4) | 0.950 | 30504(80.9) | 24764(61.0) | 0.171 |
| Number of eligible children at baseline in cluster (above median) | 12752(29.9) | 17538(42.4) | 0.773 | 17012(41.0) | 16186(33.7) | 0.886 | 25531(61.5) | 17530(52.5) | 0.574 | 29499(78.2) | 35093(86.4) | 0.603 |
| Ward of residence |  |  |  |  |  |  |  |  |  |  |  |  |
| 2 | 14592(34.2) | 19477(47.1) | Ref. | 21136(50.9) | 16139(33.6) | Ref. | 19614(47.3) | 12428(37.2) | Ref. | 7449(19.7) | 18033(44.4) | Ref. |
| 3 | 18657(43.7) | 15212(36.8) | 0.183 | 6418(15.5) | 10867(22.6) | 0.670 | 6726(16.2) | 11599(34.7) | 0.665 | 6750(17.9) | 6490(16.0) | 0.637 |
| 5 | 9406(22.1) | 6677(16.1) | 0.733 | 13944(33.6) | 21050(43.8) | 0.071 | 15150(36.5) | 9364(28.0) | 0.951 | 23529(62.4) | 16087(39.6) | 0.102 |

Data are n (%), mean ± standard deviation.

Abbreviations: JE, Japanese encephalitis vaccine; Vi-TT, Vi-tetanus toxoid conjugate vaccine.

^*^P values were generated using generalized linear mixed-effect or linear mixed-effect models, with each cluster considered as a group.

**Supplementary table 4. Baseline characteristics overall population in the two study arms stratified by vaccine coverage of under 16-year olds**

| **Characteristics** | **Coverage: 61.1% to 66.9%** | | | **Coverage: 67.0% to 69.3%** | | | **Coverage: 69.4% to 72.0%** | | | **Coverage: 72.1% to 78.9%** | | |
| --- | --- | --- | --- | --- | --- | --- | --- | --- | --- | --- | --- | --- |
|  | JE  (n=46774) | Vi-TT  (n=36064) | P^*^ | JE  (n=31485) | Vi-TT  (n=52247) | P^*^ | JE  (n=45168) | Vi-TT  (n=36804) | P^*^ | JE  (n=39944) | Vi-TT  (n=38308) | P^*^ |
| Mean age, year at start date | 25.8 ± 17.5 | 25.8 ± 17.4 | 0.812 | 25.2 ± 17.2 | 25.3 ± 17.1 | 0.695 | 25.2 ± 17.3 | 24.9 ± 17.2 | 0.225 | 25.2 ± 17.0 | 25.1 ± 17.2 | 0.577 |
| Number of male participants | 23369(50.0) | 17844(49.5) | 0.329 | 17897(49.0) | 29086(49.7) | 0.181 | 20064(50.0) | 15145(49.7) | 0.584 | 19803(49.6) | 19023(49.7) | 0.872 |
| Flush toilet facility in the house | 3356(7.2) | 2818(7.8) | 0.316 | 1548(4.2) | 3730(6.4) | 0.658 | 1097(2.7) | 1490(4.9) | 0.935 | 1803(4.5) | 769(2.0) | 0.014 |
| Own source of drinking water in the house | 22713(48.6) | 15988(44.3) | 0.367 | 12544(34.4) | 22021(37.6) | 0.858 | 13241(33.0) | 10630(34.9) | 0.474 | 14763(37.0) | 13891(36.3) | 0.771 |
| Treated drinking water | 37594(80.4) | 30291(84.0) | 0.338 | 25883(70.9) | 47141(80.5) | 0.327 | 27334(68.1) | 18457(60.5) | 0.125 | 31445(78.7) | 23640(61.7) | 0.237 |
| Handwashing after defecation | 45539(97.4) | 35542(98.6) | 0.519 | 35248(96.6) | 57774(98.7) | 0.071 | 39518(98.4) | 29483(96.7) | 0.149 | 39083(97.8) | 36615(95.6) | 0.297 |
| Longer than median distance to the midpoint of nearest treatment facility | 22017(47.1) | 15716(43.6) | 0.931 | 16647(45.6) | 37051(63.3) | 0.270 | 26629(66.3) | 15522(50.9) | 0.389 | 19216(48.1) | 15135(39.5) | 0.767 |
| Number of eligible children at baseline in cluster (above median) | 18288(39.1) | 16520(45.8) | 0.870 | 16803(46.0) | 31003(52.9) | 0.832 | 24618(61.3) | 21965(72.0) | 0.400 | 25085(62.8) | 16859(44.0) | 0.070 |
| Ward of residence |  |  |  |  |  |  |  |  |  |  |  |  |
| 2 | 10699(22.9) | 11158(30.9) | Ref. | 14623(40.1) | 19945(34.1) | Ref. | 12620(31.4) | 17272(56.6) | Ref. | 24849(62.2) | 17702(46.2) | Ref. |
| 3 | 23125(49.4) | 20271(56.2) | 0.781 | 6478(17.8) | 17569(30.0) | 0.660 | 4831(12.0) | 208(0.7) | 0.648 | 4117(10.3) | 6120(16.0) | 0.828 |
| 5 | 12950(27.7) | 4635(12.9) | 0.113 | 15391(42.2) | 21043(35.9) | 0.790 | 22710(56.5) | 13014(42.7) | 0.156 | 10978(27.5) | 14486(37.8) | 0.732 |

Data are n (%), mean ± standard deviation.

Abbreviations: JE, Japanese encephalitis vaccine; Vi-TT, Vi-tetanus toxoid conjugate vaccine.

^*^P values were generated using generalized linear mixed-effect or linear mixed-effect models, with each cluster considered as a group.

**Supplementary table 5. Baseline characteristics of total population in the two study arms stratified by vaccine coverage of the entire population**

| **Characteristic** | **Coverage: 16.8% to 18.9%** | | | **Coverage: 19.0% to 20.0%** | | | **Coverage: 20.1% to 21.4%** | | | **Coverage: 21.5% to 26.0%** | | |
| --- | --- | --- | --- | --- | --- | --- | --- | --- | --- | --- | --- | --- |
|  | JE  (n=7951) | Vi-TT  (n=7784) | P^*^ | JE  (n=7000) | Vi-TT  (n=9476) | P^*^ | JE  (n=9904) | Vi-TT  (n=7052) | P^*^ | JE  (n=8510) | Vi-TT  (n=9484) | P^*^ |
| Mean age, year at start date | 6.8 ± 4.5 | 6.7 ± 4.5 | 0.639 | 6.9 ± 4.5 | 7.0 ± 4.5 | 0.522 | 7.0 ± 4.5 | 7.1 ± 4.5 | 0.333 | 7.0 ± 4.4 | 7.2 ± 4.5 | 0.049 |
| Number of male participants | 3953(49.7) | 3910(50.2) | 0.648 | 4183(50.9) | 4709(49.7) | 0.261 | 4282(49.3) | 3470(49.2) | 0.931 | 4293(50.4) | 4725(49.8) | 0.554 |
| Flush toilet facility in the house | 529(6.7) | 512(6.6) | 0.690 | 454(5.5) | 705(7.4) | 0.850 | 199(2.3) | 252(3.6) | 0.992 | 102(1.2) | 69(0.7) | 0.349 |
| Own source of drinking water in the house | 3199(40.2) | 3284(42.2) | 0.869 | 3239(39.4) | 3832(40.4) | 0.743 | 2764(31.8) | 2264(32.1) | 0.886 | 2809(33.0) | 2645(27.9) | 0.259 |
| Treated drinking water | 6790(85.4) | 6665(85.6) | 0.644 | 6899(83.9) | 8011(84.5) | 0.793 | 5966(68.7) | 5042(71.5) | 0.429 | 4653(54.7) | 4109(43.3) | 0.403 |
| Handwashing after defecation | 7754(97.5) | 7655(98.3) | 0.603 | 8023(97.6) | 9336(98.5) | 0.543 | 8600(99.0) | 6939(98.4) | 0.695 | 8113(95.3) | 8986(94.7) | 0.950 |
| Longer than median distance to the midpoint of nearest treatment facility | 3459(43.5) | 3418(43.9) | 0.921 | 2691(32.7) | 4511(47.6) | 0.331 | 4633(53.3) | 3741(53.0) | 0.660 | 6828(80.2) | 5831(61.5) | 0.142 |
| Number of eligible children at baseline in cluster (above median) | 2391(30.1) | 3341(42.9) | 0.337 | 3347(40.7) | 3205(33.8) | 0.466 | 5288(60.9) | 3726(52.8) | 0.582 | 6668(78.4) | 8235(86.8) | 0.487 |
| Ward of residence |  |  |  |  |  |  |  |  |  |  |  |  |
| 2 | 2620(33.0) | 3578(46.0) | Ref. | 4159(50.6) | 3150(33.2) | Ref. | 4004(46.1) | 2586(36.7) | Ref. | 1713(20.1) | 4193(44.2) | Ref. |
| 3 | 3544(44.6) | 2927(37.6) | 0.498 | 1271(15.5) | 2160(22.8) | 0.770 | 1501(17.3) | 2488(35.3) | 0.622 | 1529(18.0) | 1510(15.9) | 0.554 |
| 5 | 1787(22.5) | 1279(16.4) | 0.055 | 2789(33.9) | 4166(44.0) | 0.268 | 3180(36.6) | 1978(28.0) | 0.950 | 5268(61.9) | 3781(39.9) | 0.059 |

Data are n (%), mean ± standard deviation.

Abbreviations: JE, Japanese encephalitis vaccine; Vi-TT, Vi-tetanus toxoid conjugate vaccine.

^*^P values were generated using generalized linear mixed-effect or linear mixed-effect models, with each cluster considered as a group.

**Supplementary table 6. Baseline characteristics total population in the two study arms stratified by vaccine coverage of under 16-year olds**

| **Characteristics** | **Coverage: 61.1% to 66.9%** | | | **Coverage: 67.0% to 69.3%** | | | **Coverage: 69.4% to 72.0%** | | | **Coverage: 72.1% to 78.9%** | | |
| --- | --- | --- | --- | --- | --- | --- | --- | --- | --- | --- | --- | --- |
|  | JE  (n=9026) | Vi-TT  (n=7030) | P^*^ | JE  (n=6343) | Vi-TT  (n=10462) | P^*^ | JE  (n=9643) | Vi-TT  (n=7966) | P^*^ | JE  (n=8353) | Vi-TT  (n=8338) | P^*^ |
| Mean age, year at start date | 6.9 ± 4.5 | 6.9 ± 4.5 | 0.798 | 6.9 ± 4.5 | 7.0 ± 4.5 | 0.583 | 7.0 ± 4.5 | 7.0 ± 4.5 | 0.533 | 7.0 ± 4.5 | 7.1 ± 4.5 | 0.080 |
| Number of male participants | 4538(50.3) | 3505(49.9) | 0.710 | 3652(49.5) | 5908(49.8) | 0.759 | 4325(50.3) | 3227(49.2) | 0.352 | 4196(50.2) | 4174(50.1) | 0.874 |
| Flush toilet facility in the house | 528(5.8) | 461(6.6) | 0.296 | 255(3.5) | 659(5.6) | 0.571 | 200(2.3) | 258(3.9) | 0.790 | 301(3.6) | 160(1.9) | 0.027 |
| Own source of drinking water in the house | 4117(45.6) | 2825(40.2) | 0.293 | 2326(31.5) | 4146(34.9) | 0.680 | 2647(30.8) | 2143(32.7) | 0.495 | 2921(35.0) | 2911(34.9) | 0.885 |
| Treated drinking water | 7120(78.9) | 5863(83.4) | 0.357 | 5008(67.8) | 9238(77.8) | 0.282 | 5733(66.7) | 3807(58.1) | 0.142 | 6447(77.2) | 4919(59.0) | 0.185 |
| Handwashing after defecation | 8786(97.3) | 6917(98.4) | 0.614 | 7081(95.9) | 11690(98.5) | 0.076 | 8476(98.5) | 6357(96.9) | 0.320 | 8147(97.5) | 7952(95.4) | 0.378 |
| Longer than median distance to the midpoint of nearest treatment facility | 4323(47.9) | 3099(44.1) | 0.828 | 3386(45.8) | 7643(64.4) | 0.240 | 5687(66.1) | 3439(52.4) | 0.342 | 4215(50.5) | 3320(39.8) | 0.210 |
| Number of eligible children at baseline in cluster (above median) | 3608(40.0) | 3259(46.4) | 0.693 | 3418(46.3) | 6510(54.8) | 0.791 | 5435(63.2) | 4878(74.4) | 0.493 | 5233(62.6) | 3860(46.3) | 0.289 |
| Ward of residence |  |  |  |  |  |  |  |  |  |  |  |  |
| 2 | 2117(23.5) | 2088(29.7) | Ref. | 2781(37.7) | 3897(32.8) | Ref. | 2604(30.3) | 3679(56.1) | Ref. | 4994(59.8) | 3843(46.1) | Ref. |
| 3 | 4410(48.9) | 4040(57.5) | 0.804 | 1418(19.2) | 3656(30.8) | 0.684 | 1086(12.6) | 43(0.7) | 0.523 | 931(11.1) | 1346(16.1) | 0.632 |
| 5 | 2499(27.7) | 902(12.8) | 0.041 | 3186(43.1) | 4317(36.4) | 0.802 | 4911(57.1) | 2836(43.2) | 0.193 | 2428(29.1) | 3149(37.8) | 0.784 |

Data are n (%), mean ± standard deviation.

Abbreviations: JE, Japanese encephalitis vaccine; Vi-TT, Vi-tetanus toxoid conjugate vaccine.

^*^P values were generated using generalized linear mixed-effect or linear mixed-effect models, with each cluster considered as a group.

**Supplementary table 7. Overall protection by Vi-TT vaccine**

| **Coverage (%)** | **JE arm** | | | **Vi-TT arm** | | | **Overall protection (OP)** | | | |
| --- | --- | --- | --- | --- | --- | --- | --- | --- | --- | --- |
|  | N | Typhoid  cases | Incidence rate per  100000 PY (95%CI) | N | Typhoid  cases | Incidence rate per  100000 PY (95%CI) | Crude OP  (95%CI) | P | Adjusted OP  (95%CI) | P |
| All participants | 163371 | 449 | 209(190,229) | 163423 | 206 | 96(84,110) | 54(41,65) | <0.001 | 53(40,63) ^*^ | <0.001 |
| **Entire population** |  |  |  |  |  |  |  |  |  |  |
| 16.8-18.9 | 43491 | 122 | 220(184,262) | 42473 | 66 | 125(98,158) | 44(12,64) | 0.011 | 45(22,62) ^*^ | 0.001 |
| 19.0-20.0 | 42868 | 120 | 222(185,265) | 49303 | 63 | 101(78,128) | 55(28,72) | 0.001 | 54(27,71) ^*^ | 0.001 |
| 20.1-21.4 | 42672 | 112 | 203(168,243) | 34617 | 43 | 97(71,129) | 52(12,74) | 0.019 | 45(6,68) ^*^ | 0.030 |
| 21.5-26.0 | 38449 | 95 | 189(153,229) | 41534 | 34 | 63(44,87) | 67(46,80) | <0.001 | 66(45,78) ^*^ | <0.001 |
| **<16-year** |  |  |  |  |  |  |  |  |  |  |
| 61.1-66.9 | 47678 | 131 | 208(174,246) | 37264 | 49 | 105(79,138) | 50(8,73) | 0.025 | 49(19,67) ^*^ | 0.004 |
| 67.0-69.3 | 37397 | 120 | 256(213,305) | 60259 | 81 | 109(87,135) | 58(36,72) | <0.001 | 54(32,68) ^*^ | <0.001 |
| 69.4-72.0 | 41522 | 109 | 201(166,242) | 31719 | 30 | 73(50,102) | 64(37,80) | <0.001 | 62(34,79) ^*^ | 0.001 |
| 72.1-78.9 | 40898 | 89 | 175(142,215) | 39513 | 46 | 89(66,117) | 48(16,68) | 0.008 | 47(10,69) ^†^ | 0.019 |

Abbreviations: JE, Japanese encephalitis vaccine; Vi-TT, Vi-tetanus toxoid conjugate vaccine; N, number of participants; PY, person years; CI, confidence interval.

^*^Adjusted for geographical ward, distance to study clinics, number of eligible children.

^†^Adjusted for flush toilet in the house, geographical ward, distance to study clinics, number of eligible children.

**Supplementary table 8. Total protection by Vi-TT vaccine**

| **Coverage (%)** | **JE arm** | | | **Vi-TT arm** | | | **Total protection (TP)** | | | |
| --- | --- | --- | --- | --- | --- | --- | --- | --- | --- | --- |
|  | N | Typhoid  cases | Incidence rate per  100000 PY (95%CI) | N | Typhoid  cases | Incidence rate per  100000 PY (95%CI) | Crude TP  (95%CI) | P | Adjusted TP  (95%CI) | P |
| All participants | 33365 | 264 | 610(540,687) | 33796 | 45 | 103(76,137) | 83(74,89) | <0.001 | 83(74,89) ^*^ | <0.001 |
| **Entire population** |  |  |  |  |  |  |  |  |  |  |
| 16.8-18.9 | 8153 | 75 | 756(599,942) | 8053 | 14 | 145(83,237) | 81(49,93) | 0.001 | 82(59,92) ^*^ | <0.001 |
| 19.0-20.0 | 8587 | 69 | 652(511,820) | 9799 | 14 | 114(66,187) | 82(67,91) | <0.001 | 81(66,90) ^*^ | <0.001 |
| 20.1-21.4 | 8993 | 68 | 593(465,747) | 7385 | 8 | 86(41,163) | 85(68,93) | <0.001 | 83(64,91) ^*^ | <0.001 |
| 21.5-26.0 | 8701 | 52 | 460(348,599) | 9689 | 9 | 72(35,131) | 84(66,93) | <0.001 | 82(60,92) ^†^ | <0.001 |
| **<16-year** |  |  |  |  |  |  |  |  |  |  |
| 61.1-66.9 | 9260 | 77 | 656(521,815) | 7332 | 10 | 115(59,205) | 82(32,95) | 0.012 | 82(50,94) ^*^ | 0.012 |
| 67.0-69.3 | 7586 | 73 | 786(621,983) | 12295 | 18 | 121(74,188) | 85(73,91) | <0.001 | 84(73,90) ^*^ | <0.001 |
| 69.4-72.0 | 8993 | 59 | 516(397,661) | 6873 | 6 | 68(28,140) | 87(71,94) | <0.001 | 86(70,94) ^*^ | <0.001 |
| 72.1-78.9 | 8593 | 55 | 509(387,657) | 8638 | 11 | 97(52,168) | 81(61,90) | <0.001 | 80(60,90) ^‡^ | <0.001 |

Abbreviations: JE, Japanese encephalitis vaccine; Vi-TT, Vi-tetanus toxoid conjugate vaccine; N, number of participants; PY, person years; CI, confidence interval.

^*^Adjusted for geographical ward, distance to study clinics, number of eligible children.

^†^Adjusted for age, geographical ward, distance to study clinics, number of eligible children.

^‡^Adjusted for flush toilet in the house, treated drinking water, geographical ward, distance to study clinics, number of eligible children.

**Supplementary table 9. Overall protection by Vi-TT vaccine among participants under 16 years of age**

| **Coverage (%)** | **JE arm** | | | **Vi-TT arm** | | | **Overall protection (OP)** | | | |
| --- | --- | --- | --- | --- | --- | --- | --- | --- | --- | --- |
|  | N | Typhoid  cases | Incidence rate per  100000 PY (95%CI) | N | Typhoid  cases | Incidence rate per  100000 PY (95%CI) | Crude OP  (95%CI) | P | Adjusted OP  (95%CI) | P |
| **All participants** | 51878 | 355 | 518(466,574) | 52333 | 131 | 190(160,225) | 63(50,73) | <0.001 | 62(50,72) ^*^ | <0.001 |
| **Entire population** |  |  |  |  |  |  |  |  |  |  |
| 16.8-18.9 | 13046 | 102 | 616(505,744) | 12922 | 37 | 229(164,312) | 63(34,79) | 0.001 | 65(44,78) ^*^ | 0.001 |
| 19.0-20.0 | 13520 | 97 | 569(464,691) | 15231 | 37 | 190(136,260) | 67(43,81) | <0.001 | 65(43,79) ^*^ | <0.001 |
| 20.1-21.4 | 13773 | 88 | 495(399,606) | 11238 | 35 | 243(172,334) | 51(9,74) | 0.024 | 43(2,67) ^*^ | 0.044 |
| 21.5-26.0 | 12995 | 68 | 397(311,500) | 14508 | 22 | 117(75,173) | 71(50,83) | <0.001 | 68(47,81) ^*^ | <0.001 |
| **<16-year** |  |  |  |  |  |  |  |  |  |  |
| 61.1-66.9 | 14998 | 108 | 544(449,654) | 11686 | 30 | 204(141,287) | 63(22,82) | 0.008 | 61(32,78) ^*^ | 0.001 |
| 67.0-69.3 | 11926 | 94 | 627(510,764) | 19251 | 47 | 198(148,261) | 69(48,81) | <0.001 | 64(43,78) ^*^ | <0.001 |
| 69.4-72.0 | 13564 | 88 | 498(402,610) | 10557 | 20 | 145(92,220) | 71(52,82) | <0.001 | 70(51,82) ^*^ | <0.001 |
| 72.1-78.9 | 12826 | 65 | 406(316,514) | 12727 | 34 | 203(143,281) | 49(10,71) | 0.021 | 51(8,74) ^†^ | 0.025 |

Abbreviations: JE, Japanese encephalitis vaccine; Vi-TT, Vi-tetanus toxoid conjugate vaccine; N, number of participants; PY, person years; CI, confidence interval.

^*^Adjusted for geographical ward, distance to study clinics, number of eligible children.

^†^Adjusted for flush toilet in the house, geographical ward, distance to study clinics, number of eligible children.
